# Clinical management and impact of scarlet fever in the modern era: findings from a cross-sectional study of cases in London, 2018-2019

**DOI:** 10.1101/2021.06.25.21259535

**Authors:** M. Trent Herdman, Rebecca Cordery, Basel Karo, Amrit Kaur Purba, Lipi Begum, Theresa Lamagni, Chuin Kee, Sooria Balasegaram, Shiranee Sriskandan

## Abstract

**Background:** Scarlet fever incidence has risen steeply in recent years, and is associated with wider outbreaks of severe Group A Streptococcal infections. Yet, few studies of its epidemiology, clinical features, and management have been undertaken in the antibiotic era.

**Aim:** To characterize symptomatology, management, and consequences of scarlet fever and identify associations with delayed diagnosis.

**Design/Setting:** Cross-sectional study of children with scarlet fever in London, 2018-2019.

**Methods:** online survey of parents/guardians of children with scarlet fever identified by Health Protection Teams, recording demographics, symptoms, care-seeking, and management; logistic regression for factors associated with delayed diagnosis; Cox’s regression for consequences of delayed diagnosis.

**Results:** Responses represented 412 cases in a period with 6828 notifications for children 0-14 years old, and 410 school/nursery outbreaks. 70% first sought care from general practice, and 31% had multiple consultations. For 28%, scarlet fever was not considered at first consultation: in these cases, symptoms were frequently attributed to viral infection (60%, 64/106). Delay in diagnosis beyond first consultation occurred more frequently among children aged 5+ who presented with sore throat (odds ratio 2.8 *vs*. 5+ without sore throat; 95%CI 1.3-5.8; P=0.006). On average, cases with delayed diagnosis took one day longer to return to baseline activities, and required one additional day off school *versus* those diagnosed at first consultation.

**Conclusions:** In assessing children with fever, rash, and sore throat, practitioners should be alert to the possibility of scarlet fever: it is frequently missed at first consultation, and prompt recognition speeds clinical recovery and public health management.

**How this fits in [4 sentences summarising key messages of background and findings]:**

- In the five years leading up to the pandemic lockdown of 2020, scarlet fever incidence rose markedly in England and Wales, prompting this investigation of cases in London 2018-2019.
- Prompt recognition of the disease by carers and clinicians can direct timely antibiotic therapy, limit transmission in the household and community, and direct the identification and control of outbreaks.
- In this study, delayed diagnosis was more likely to occur among older children presenting with sore throat—perhaps reflecting a lower index of suspicion in this age group.
- Cases with delayed diagnosis took longer to return to normal activities, and required more time off from school than those diagnosed at the first consultation.

## Introduction

### Background and Rationale

Scarlet fever results from pharyngeal infection with group A streptococci (GAS) expressing erythrogenic toxins, classically leading to rash, fever, constitutional symptoms, and localized symptoms including sore throat(1). Most studies of scarlet fever predate a modern understanding of streptococcal infections and their treatment(2-4). Incidence of scarlet fever in England and Wales declined from the 1940s to the mid-2010s, but increased markedly in the five years leading up to the pandemic lockdown of 2020: this coincided with the emergence of a dominant, more toxigenic lineage of GAS(5-7). Given the association between scarlet fever and severe GAS infections, including invasive soft tissue infections and toxic shock syndrome, this upsurge in cases underlined the need to review clinical practice(5, 8-10).

Scarlet fever is a notifiable infectious disease in England, and usually diagnosed on the basis of symptoms and signs. The triad of rash, sore throat, and fever is typical, but this presentation may be mistaken for viral infection, delaying the administration of antibiotics(11). Early case recognition aids implementation of treatment, initiation of which should reduce likelihood of complications and prevent onward transmission, reducing the incidence of not only scarlet fever itself, but also invasive GAS infections in cases and contacts(10).

### Objectives

Prompted by increased scarlet fever notification, we undertook a survey of scarlet fever cases in London, to characterise the presenting features and healthcare experience, focussing on the timing of diagnosis and interventions, and impact on cases and their households(10, 12).

## Methods

### Study Design and Recruitment

We conducted a cross-sectional, observational study of confirmed or probable scarlet fever cases using an online survey (SelectSurvey v.4.0, **Supplementary File 1**). Invitations were sent to parents and guardians of all cases of scarlet fever notified by clinicians to Public Health England (PHE) Health Protection Teams (HPTs) in March-May 2018 in London. In March-May 2019, a modified version of the survey (omitting or rewording some questions, adding others) was sent to parents/guardians of notified sporadic cases plus cases identified as part of notified school outbreaks (**Supplementary File 2**). Participation was voluntary, anonymised, and approved by a national Research Ethics Committee (London-Chelsea REC Reference 18/LO/0025; IRAS Reference 225006). Consent was inferred from survey participation.

Public health management of cases and outbreaks was according to national guidelines used by HPTs(12).

### Data Analysis

Quantitative data description and analysis were performed using Stata14.2 and GraphPad Prism7.0 (see **Supplementary Methods** for further details). Ethnicity proportions were compared to Department for Education primary schools data for London(13). In assessing symptoms and signs, description was restricted to cases diagnosed by a health professional.

To identify variables associated with delayed diagnosis, we compared cases where scarlet fever was suspected at first contact with a health professional versus those for whom it was not, restricting to cases formally notified by a health professional. A logistic regression model was constructed using a stepwise, subtractive approach. Models were compared using Akaike’s information criteria and Bayesian information criteria, with likelihood ratio tests used to address ambiguous comparisons, and stratifying as required to address effect modification.

Consequences of delayed diagnosis were assessed in terms of days until recovery to normal activity, days of school/nursery missed, and days of work missed by carers, constructing Cox’s proportional hazards regression models (condition of proportionality was met). The model for time to recovery to normal activity was limited to data from the 2019 survey, as it was not ascertained in 2018.

### Qualitative textual analysis

Free text volunteered by respondents was coded in NVivo13 and Microsoft Excel using a thematic matrix for responses concerning perception of scarlet fever, and analysed to characterize experiences of the illness, accessibility of information and care, experiences of the health service, and impact on the case and their household, and identify ramifications for providers of clinical practice and health protection.

## Results

### Baseline characteristics

London HPTs identified 4172 cases of confirmed or probable scarlet fever in children 0-14 years old, and 263 school/nursery outbreaks in 2018, and 2656 cases and 147 school/nursery outbreaks in 2019. From 1 March-31 May 2018, we contacted parents or guardians of 1703 cases of scarlet fever notified to London HPTs. From 1 March-31 May 2019 recruitment took place both through HPT contact with 872 identified scarlet fever cases and through dissemination to parental networks by outbreak-affected schools/nurseries. Surveys were completed for 477 children, 412 of whom met the case definition (339 in 2018, and 73 in 2019). In 381 cases (92%), scarlet fever was diagnosed by a health professional; 31 cases (8%) had a confirmed epidemiological link to an outbreak, but may not have been diagnosed by a health professional, and hence were excluded from analyses of clinical features.

Demographic and baseline clinical characteristics are described in **Table 1**. Compared to 2015 Department for Education estimates for primary schools in London, responses showed a higher proportion of White participants (70% vs. 42% in primary schools, P<0.001) and lower proportions of participants of Asian/Asian British (11% vs. 20%, P<0.001) and Black/African/Caribbean/Black British ethnicity (5% vs. 21%, P<0.001) (13).

**Table 1:** Demographic characteristics of participating scarlet fever cases (n=412).

| Baseline Characteristics | All Cases |  |
| --- | --- | --- |
|  | N | (%) |
| **Age Group** |  |  |
| 0-2 years | 66 | (16%) |
| 3-4 years | 156 | (38%) |
| 5-9 years | 177 | (43%) |
| 10-16 years | 12 | (3%) |
| *missing* | 1 | . |
| **Sex** |  |  |
| Female | 197 | (48%) |
| Male | 212 | (51%) |
| *missing/prefer not to say* | 3 | . |
| **Ethnicity** |  |  |
| Asian/Asian British | 47 | (11%) |
| Black/African/Caribbean/Black British | 21 | (5%) |
| Mixed/Multiple Ethnicities | 48 | (12%) |
| White | 287 | (70%) |
| Other | 5 | (1%) |
| *missing/prefer not to say* | 4 | . |
| **School Group** |  |  |
| Nursery/Playgroup | 167 | (41%) |
| Reception Class | 85 | (21%) |
| Primary School Year 1 | 48 | (12%) |
| Primary School Year 2 | 31 | (8%) |
| Primary School Year 3 | 28 | (7%) |
| School Beyond Year 3 | 33 | (8%) |
| *Missing/none volunteered* | 20 | . |
| **General health prior to scarlet fever** |  |  |
| Ever hospitalized | 107 | (26%) |
| Follow-up in out-patient clinic | 38 | (9%) |
| Chronic underlying illness<sup>1</sup> | 8 | (2%) |
| **Upper respiratory tract history** |  |  |
| ≥1 episode of sore throat in preceding year | 190 | (46%) |
| Previous isolation of GAS | 13 | (3%) |
| Previous tonsillectomy | 11 | (3%) |

<sup>1</sup>4 report asthma; 3 report recurrent tonsillitis.

### Clinical characteristics

Rash was the most commonly identified symptom, reported by 89% of respondents (**Table 2**). However, fever and sore throat were more likely than rash to be noted first. Among respondents commenting on the timing of the rash relative to other symptoms, 71% (32 of 45 responding) reported the rash followed other symptoms, with a median one-day delay (IQR 0-2.5 days; range 0-15 days). Cases with a history of recurrent sore throat were more likely to present with sore throat initially (OR 1.9, 95%CI 1.2-2.9, P=0.008), and substantially more likely to experience a sore throat at some point in the illness (OR 11.3, 95%CI 4.4-29.3, P<0.001).

**Table 2:** Reported Symptoms among 381 cases diagnosed with scarlet fever by a health professional within 4 weeks of survey completion.

| Symptom | Under 5 years old |  | 5 years and older |  | All ages |  |
| --- | --- | --- | --- | --- | --- | --- |
|  | n/total | (%) | n/total | (%) | n/total | (%) |
| **First Symptom(s) Noted:** |  |  |  |  |  |  |
| Fever | 97/180 | (54%) | 57/158 | (36%) | 154/338 | (46%) |
| Sore Throat | 48/180 | (27%) | 84/158 | (53%) | 132/338 | (39%) |
| Rash | 72/180 | (40%) | 43/158 | (27%) | 115/338 | (34%) |
| Not Playing/Tiredness | 36/180 | (20%) | 21/158 | (13%) | 57/338 | (17%) |
| **Symptom Ever Noted:** |  |  |  |  |  |  |
| Rash | 187/207 | (90%) | 149/170 | (88%) | 336/377 | (89%) |
| Fever | 184/204 | (90%) | 143/166 | (86%) | 327/370 | (88%) |
| Sore Throat | 151/192 | (79%) | 138/163 | (85%) | 289/355 | (81%) |
| Tiredness | 136/180 | (76%) | 113/158 | (72%) | 249/338 | (73%) |
| Enlarged Tonsils | 100/155 | (65%) | 80/124 | (65%) | 180/279 | (65%) |
| Not Eating | 126/180 | (70%) | 90/158 | (57%) | 216/338 | (64%) |
| Not Playing | 86/180 | (48%) | 72/158 | (46%) | 158/338 | (47%) |
| Headache | 53/180 | (29%) | 71/158 | (45%) | 124/338 | (37%) |
| Pus on Tonsils | 54/151 | (36%) | 47/119 | (40%) | 101/270 | (37%) |
| Sore Tongue | 57/180 | (32%) | 45/158 | (28%) | 102/338 | (30%) |
| Stomach Ache | 44/180 | (24%) | 50/158 | (32%) | 94/338 | (28%) |
| Vomiting | 48/180 | (27%) | 29/158 | (18%) | 77/338 | (23%) |
| Swollen Tongue | 28/180 | (16%) | 24/158 | (15%) | 52/338 | (15%) |
| Earache | 21/180 | (12%) | 29/158 | (18%) | 50/338 | (15%) |
| Diarrhoea | 24/180 | (13%) | 14/158 | (9%) | 38/338 | (11%) |

70% of respondents characterizing the rash (19/27) described it as sand-papery or rough to feel, 63% (17/27) as red, 26% (7/27) as comprising small spots, 19% (5/27) as pink, 15% (4/27) as itchy, and 4% (1/27) as peeling off. Median duration of the rash was 5 days (IQR 3-8 days; range 1-14 days). 69% (18/26) reported the rash first appeared on the trunk, 19% (5/26) on the face, and 12% (4/26) on the arms and legs. Rash was seen in 89% of White cases and 90% of cases of other ethnicities (P=0.75).

While most children (71%) eventually experienced fever, rash, and sore throat, the pattern of symptoms at onset varied with age (**Table 2**). Sore throat was a more common initial symptom among older cases (OR 3.1, 95%CI 1.9-5.0, P<0.001). Conversely, rash and fever were less likely at onset among older cases (respectively OR 0.6, 95%CI 0.4-0.9, P=0.014; OR 0.5, 95%CI 0.3-0.8, P=0.001).

### Differential diagnosis and clinical management

Table 3 summarizes the sources of care sought for cases. Median duration from onset of symptoms to seeing a health professional was 2 days (IQR 1-3 days; range<1-14 days). For 31% of cases, additional consultations were undertaken (with 14% requiring three or more consultations).

At the first consultation with a doctor, 72% of cases (268/374) had scarlet fever as the diagnosis (or part of the differential diagnosis). When the diagnosis was delayed, 60% (64/106) had their illness ascribed to a viral infection, 21% (22/106) to tonsillitis, and 13% (14/106) to pharyngitis. Throat swabs were taken from 44% of cases (148/338). Of those who knew the results of the swab, 91% (75/82) reported GAS was isolated. Antibiotic prescribing practices are described in **Supplementary Table S1**: 93% of cases were prescribed an agent consistent with clinical guidelines.

**Table 3:**
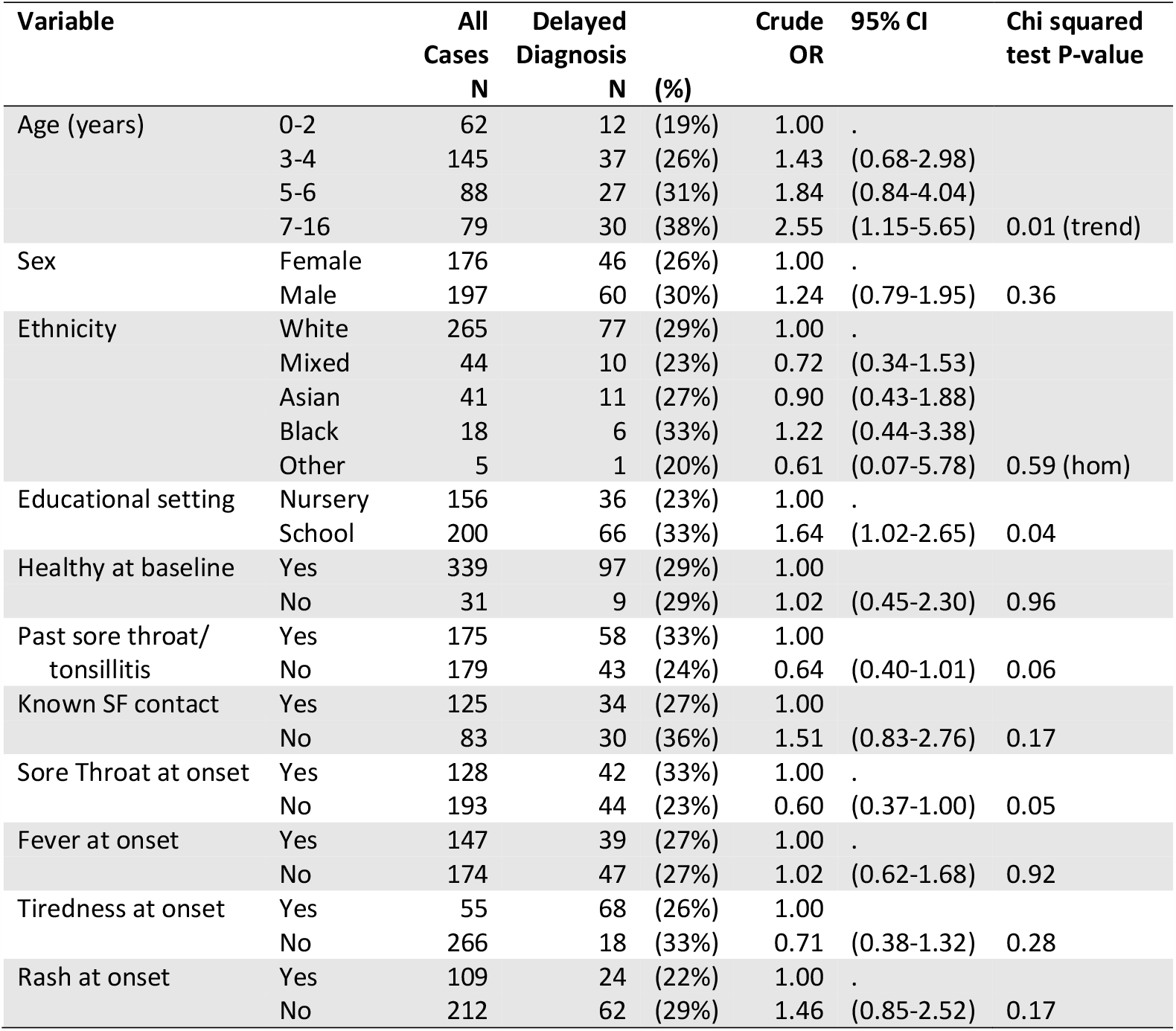
Crude analysis of demographic and clinical variables associated with delayed diagnosis (diagnosis of scarlet fever not considered at first consultation with healthcare; n=374):

### Burden and impact of disease

80% of cases (329/402) missed school because of their illness, with a median of 3 days lost (IQR 2-4 days; range 1-14 days). Median time from starting antibiotics to return to normal activity such as attending school or nursery was 2 days (IQR 1-4 days; range 0-8 days, asked only in 2019, with 71 respondents).

For 53% of cases (198/372), at least one carer took time off work, with a median total of 2 days taken as leave (IQR 1-3; range 0-11 days). In 23% of cases (92/398), a carer became ill themselves. In 22% (67/301), the child’s usual carers required additional help with care during the illness— provided by family members for 80%, friends for 5%, and paid professionals for 15%. In 11% of cases (37/337), other children in the household also missed school—predominantly because they were unwell themselves; less frequently because of dependence on the caregiver to transport siblings to school. In 2019, 43% (34/79) reported other unwell family members: 29 with sore throat, 10 with tonsillitis, 6 with scarlet fever, one each with cellulitis and conjunctivitis (11 households identified multiple illnesses).

### Risk factors for delayed diagnosis

In a logistic model for delayed diagnosis among 321 cases in the 2018 survey, the strongest fit was provided by variables for age (under 5 years vs. 5 and older), sore throat at onset, and interaction between these variables (**Table 4, Supplementary Table S1**). No other variables affected the model. Among cases aged 5 and older, those with sore throat present at symptom onset had 2.7 times the odds of a delayed diagnosis compared to those without (95%CI 1.3-5.8, P<0.01). Among cases aged under 5, we found no evidence of an association between sore throat and delayed diagnosis (aOR 0.6, 95%CI 0.3-1.5, P=0.33).

**Table 4:** Pathways of care for participating cases.

| Care Pathways (among n responding) | n | (%) |
| --- | --- | --- |
| **First Source of Advice (332):** |  |  |
| GP | 267 | (70%) |
| NHS Direct | 39 | (10%) |
| Walk-In Centre | 30 | (8%) |
| Hospital Emergency Department | 27 | (7%) |
| Internet | 26 | (7%) |
| Urgent Care Centre | 16 | (4%) |
| Local Pharmacy | 14 | (4%) |
| School Nurse | 3 | (1%) |
| **Initial Differential Included SF (367)** | 265 | (72%) |
| **Repeat visit to HCW Needed (380)** | 116 | (31%) |
| **Source of Second Consultation (116):** |  |  |
| GP | 71 | (61%) |
| Emergency Department | 14 | (12%) |
| Urgent Care Centre | 11 | (9%) |
| Other | 5 | (4%) |
| **Reason for Second Consultation (116):** |  |  |
| Child developed a new symptom(s) | 44 | (38%) |
| Worried that it could be Scarlet Fever | 37 | (32%) |
| Asked to come back if not better | 19 | (17%) |
| Child could not take prescribed medication | 6 | (5%) |
| Called back due to swab result | 6 | (5%) |
| Other* | 4 | (4%) |
| **Hospitalized (326)** | 7 | (2%) |

\*Two for further investigations, two for specialist consultation.

### Consequences of delayed diagnosis

Cases returned to normal activity faster when scarlet fever was considered at the first consultation (33/52; ascertained in 2019 only), with a median recovery time of 2 days from starting antibiotics when scarlet fever was considered, vs. 3 when it was not, and a hazard ratio (HR) for recovery of 0.53 (95%CI 0.28-0.99; P=0.047; **Supplementary Figure S1**). Cases diagnosed without delay returned to school sooner, with a median of 2 days off (246/298) and 3 days for those with delay (92/298) (HR 0.77, 95%CI 0.59-0.99; P=0.045). We found no difference in days of work missed by carers between the two groups, with a median of 2 days missed for both (HR 0.91, 95%CI 0.64-1. 29; P=0.592).

### Qualitative synthesis

In thematic analysis of 194 free-text comments (**Table 5**), some respondents reported reassurance that a diagnosis was made promptly by practitioners who recognised the syndrome: others were disappointed that antibiotic treatment was delayed where symptoms were attributed to viral infection. Representativeness of online resources was questioned, such as the difficulty in finding depictions of the rash on non-white skin. While some respondents noted rapid recovery and minimal impact, others recorded spread of streptococcal infections to carers and other household members, and a wider impact of the time demands and stress of providing care to unwell children.

**Table 5:**
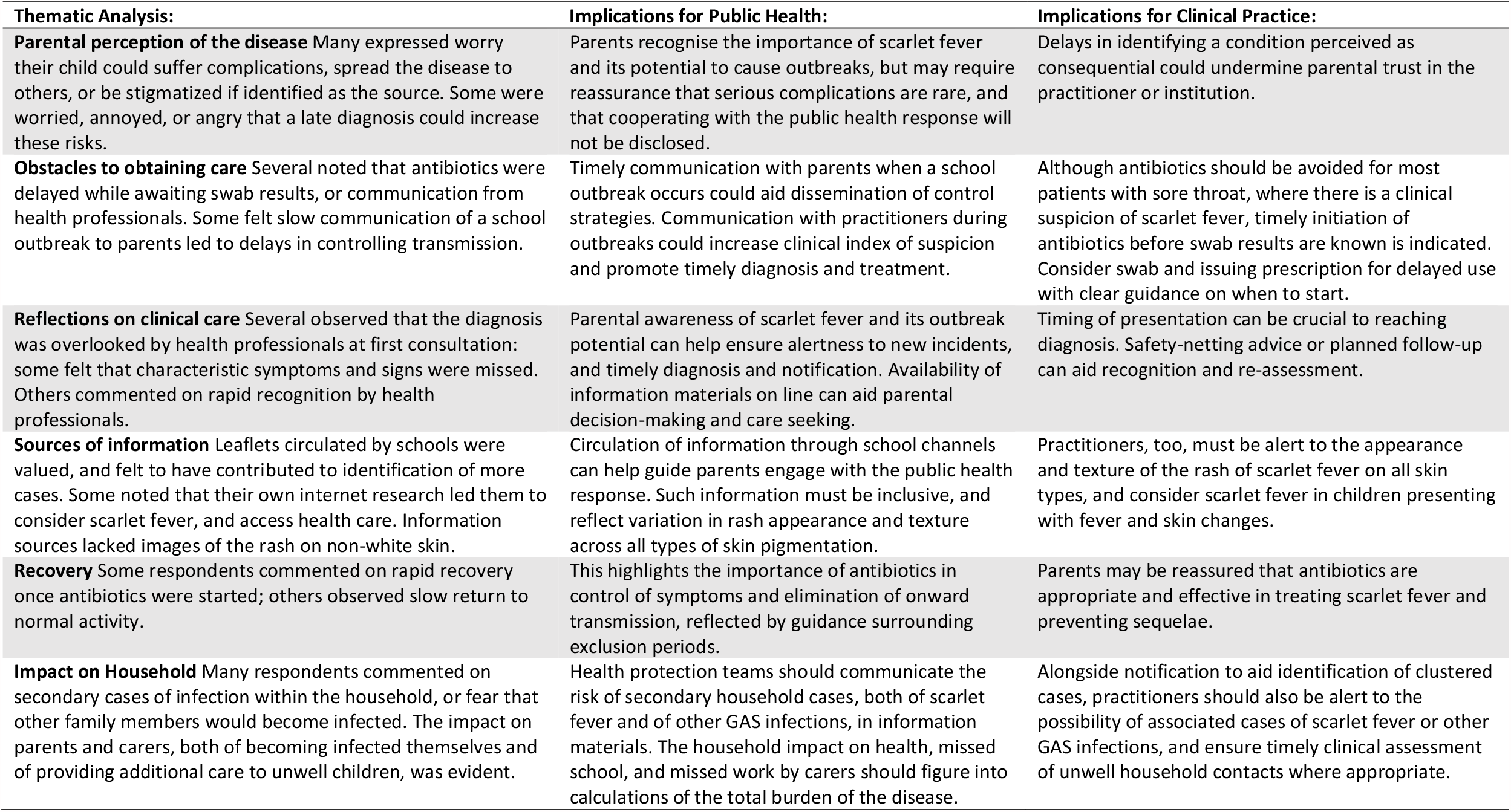
Thematic analysis of free text comments from respondents to questionnaires, 2018-2019.

## Discussion

### Summary

Undertaken at a time of increased incidence, this study provides an important update on the impact of scarlet fever, identifies opportunities for improved recognition, and highlights the previously unquantified burden of disease on affected households.

Practitioners should be alert to circumstances in which scarlet fever is easily overlooked. In this survey, a delay in diagnosis among older children was 2.8 times as likely when a sore throat was present at onset, with symptoms often ascribed to viral infection. Practitioners may have a lower index of suspicion in this age group, and be less likely to consider the diagnosis at first presentation when another explanation for symptoms is available. Timely recognition of scarlet fever in this age group could expedite antibiotic treatment, shorten the period of infectivity, and reduce onward propagation of GAS.

The sand-papery rash of scarlet fever was perceived by most carers, and tended to appear after other symptoms (median one day later). Rash timing is important in distinguishing scarlet fever from measles and rubella—which have a longer lag—but can lead to confusion with other viral exanthems(14). Awareness of the timing and sand-papery character of scarlet fever’s rash may help practitioners make the diagnosis and commence treatment.

### Comparison with existing literature

Current clinical guidance for sore throat advises primary care physicians to give antibiotics only when a more serious condition (such as suppurative infection or sepsis) is suspected(15). FeverPAIN and Centor scores are validated in rapid appraisal for GAS pharyngitis, but scarlet fever falls outside their scope(16, 17). The rash of scarlet fever—particularly during the season of increased risk from March to May—should also prompt practitioners to commence antibiotics(18). The public health importance of prompt diagnosis and treatment is underscored by the 12-fold greater risk of invasive GAS among scarlet fever household contacts(10). Advice to avoid unnecessary antibiotics for most sore throats is valuable to antimicrobial stewardship: the caveat is that scarlet fever and other GAS infections require antibiotics to prevent complications and reduce onward spread.

In this study, 80% of children missed school/nursery for a median of 3 days. Time to recovery and return to school was longer when diagnosis was delayed. As the average primary school pupil misses 7.4 days a year, this increase is substantial(19). Scarlet fever affected almost 32,000 children in the UK in 2018(20); the direct medical costs, including hospital admissions (1 in 40 case in 2014), plus the risk of secondary GAS infections, and the non-medical costs of childcare and lost education, amount to a sizeable health and economic burden(5, 21).

### Strengths and Limitations

By surveying notified scarlet fever cases, this study draws on the experience of patients and households accessing primary care. However, our use of an online survey tool introduces a risk of selection bias. Compared to the population at risk, more cases were white than would be expected by chance. There may be bias in recognition or notification, given that invasive GAS infection is observed with higher incidence in ethnicities other than white (22-24). Respondents’ observed difficulty finding illustrations of the rash on non-white skin corroborates under-representation in educational materials noted elsewhere(25-27). It also highlights the importance of ascertaining ethnicity in general practice research and surveillance, so that outbreaks affecting specific communities—or obstacles accessing care—are identified(28).

### Implications for Practice and Research

Differentiating scarlet fever from viral infections presents a clinical challenge: sore throat is common to both conditions, and the rash of scarlet fever, though characteristic, may be subtle or delayed. Point-of-care tests may be useful to avoid unnecessary antibiotic prescription. Alertness to periodic peaks in scarlet fever—from March to May in the UK—and the occurrence of local outbreaks may help set an appropriate index of suspicion(5). Increased local incidence should drive more communication with carers about symptoms of concern (such as a sand-papery rash). The need for sound antimicrobial stewardship should not preclude access to timely clinical diagnosis of scarlet fever, microbiological testing, and empirical prescribing.

The strains of GAS that cause scarlet fever also trigger outbreaks of pharyngitis and invasive GAS infections. As such, a single case of scarlet fever may signal a larger outbreak of unrecognised GAS infections(5, 7, 29). Further research into the interplay of scarlet fever and invasive GAS at a population level will help direct diagnostic and treatment strategies to reduce the impact of such outbreaks.

Meanwhile, effective control of scarlet fever and GAS depends upon the coordinated efforts of clinicians and public health practitioners to identify cases and outbreaks early, implement appropriate treatment, and prevent onward transmission.

## Supporting information

Supplementary File 1: Questionnaire 2018

Supplementary File 2: Questionnaire 2019

Supplementary File 3: STROBE Checklist

Disclosure Forms

## Data Availability

Non-identifiable data used in this study will be made available to researchers upon reasonable request to the corresponding author.

## Acknowledgements

We are grateful to carers for taking time to complete the survey, and to PHE London Health Protection Teams’ Surveillance Officers for scarlet fever surveillance and data analysis. We would like to thank the staff of the London Health Protection Teams and the Field Epidemiology Service for their support with the study. SS Acknowledges the NIHR Biomedical Research Centre grant awarded to Imperial College.

## Funding

This report was funded by Action Medical Research. It was also funded in part by the Medical Research Council (grant MR/P022669/1) and the National Institute for Health Research Health Protection Research Unit (NIHR HPRU) in Healthcare Associated Infections and Antimicrobial Resistance at Imperial College London in partnership with Public Health England (grant HPRU-2012-10047). The views expressed are those of the authors and not necessarily those of the NHS, the NIHR, the Department of Health or Public Health England.

## Conflicts of Interest

The authors declare no conflicts of interest.

## Online Supplementary Material

**Supplementary File 1: 2018 Online Questionnaire**

**Supplementary File 2: 2019 Online Questionnaire**

**Supplementary File 3: STROBE Checklist for Cross-Sectional Analysis**

## Supplementary Methods for Cross-Sectional Analysis

### Participants

Children under 16 with scarlet fever were eligible if notified to London HPTs as confirmed or probable scarlet fever from March 1 to May 31 2018, or March 1 to May 31 2019. In 2018, the parents or guardians of all eligible cases for whom contact details and notification by a clinician could be confirmed were invited for participation; in 2019, parents or guardians were invited for participation after either notification by a clinician or via invitations circulated to parental networks of schools or nurseries with notified outbreaks.

The case definition of scarlet fever was in keeping with PHE guidance definitions of confirmed or probable scarlet fever: for sporadic cases identified through statutory notification (including the index cases of suspected outbreaks), a case constituted a clinical diagnosis of scarlet fever by a health professional (with or without detection of Group A Streptococci on a throat swab); in the context of an established outbreak, cases required a credible report of signs or symptoms consistent with scarlet fever with a close epidemiological link to a confirmed or probable case (with or without confirmation by a health professional).^1^

### Variables Ascertained

Surveys collected data on demographics, medical history, contact history, symptoms, care-seeking behaviour, diagnoses and clinical management by health professionals, impact upon household caregivers, and knowledge and attitudes regarding scarlet fever on the part of the responding parent or guardian. Survey texts can be found in **Supplementary Files S1 and S2**.

### Data Sources and Measurement

For analysis of variables associated with delayed diagnosis, the outcome was defined dichotomously as a case for whom scarlet fever was not considered in the differential diagnosis at the first consultation with a clinician (in the recollection of the responding parent or guardian). For analysis of consequences of delayed diagnosis, this dichotomous variable was considered as an exposure, and assessed along with potential confounders and effect modifiers for association with time to resumption of normal activity, time to return to school, and time to parent/guardian’s return to work.

### Study Size

The study size was determined pragmatically, attempting to contact as many notified cases and schools/nurseries as possible over the course of two high-transmission seasons.

### Quantitative Variables

Age in years was collected as a continuous variable and stratified for multivariable analysis into under 5 and 5 and older to increase statistical power and reflect the age at which school attendance starts. Other demographic and clinical exposure variables were ascertained and analysed dichotomously. Recovery outcomes were ascertained as continuous variables in days in the Cox’s proportional hazards regression model.

### Missing Data

Missing values were addressed in regression models by introducing an additional category for unknown values of categorical variables. Cases with missing values for the outcome variables in Cox’s regression were excluded from the analysis.

**Supplementary Table S1:** Antibiotic prescribing patterns described by respondents (n=339)

| **Antibiotic Treatment Characteristics** | **n** | **(%)** |
| --- | --- | --- |
| **Antibiotic Prescribed (n=311)** |  |  |
| Penicillin V<sup>1</sup> | 235 | (76%) |
| Amoxicillin<sup>2</sup> | 44 | (14%) |
| Azithromycin<sup>3</sup> | 15 | (5%) |
| Erythromycin | 14 | (5%) |
| Other agent | 6 | (2%) |
| **Recommended Start (n=319)** |  |  |
| Immediate (once prescribed) | 303 | (95%) |
| In the event of worsening symptoms | 16 | (5%) |
| **Prescribed Recommended Duration (n=288)** |  |  |
| Yes | 238 | (83%) |
| No | 50 | (17%) |
| **Took Full Prescribed Course (n=339)** |  |  |
| Yes | 294 | (87%) |
| No | 45 | (13%) |
| **Reason for Stopping Early (n=39)** |  |  |
| Clinical improvement | 16 | (41%) |
| Doses excessive | 7 | (18%) |
| Unpleasant taste | 6 | (15%) |
| Other reasons | 10 | (26%) |

<sup>1</sup>As recommended first line in NICE guidance; <sup>2</sup>As recommended if unable to swallow tablets; <sup>3</sup>As recommended if penicillin-allergic.

**Supplementary Table S2:**
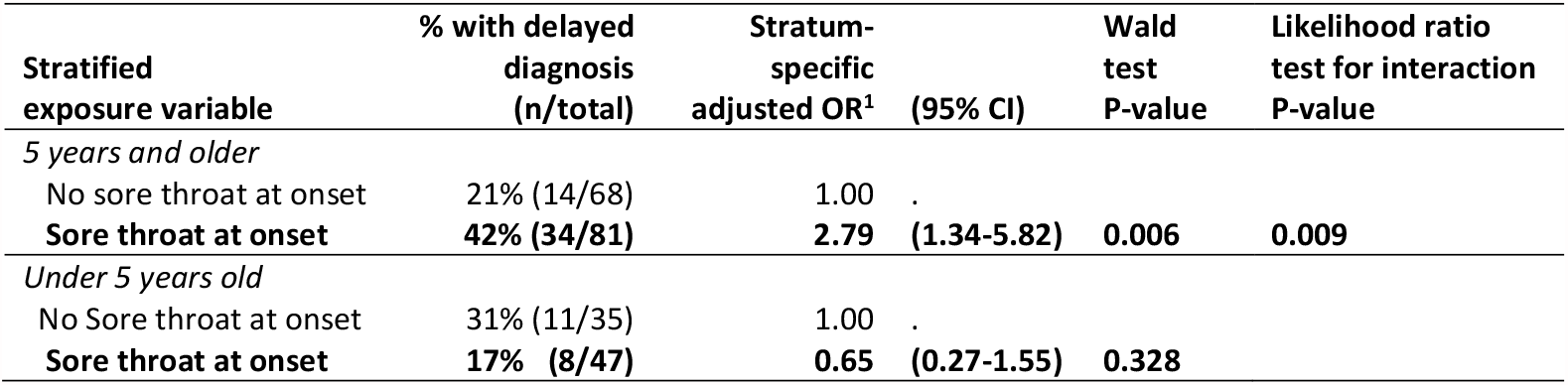
Stratified model for associations with delayed diagnosis of scarlet fever among cases in 2018 (n=321).

| **Stratified exposure variable** | **% with delayed diagnosis (n/total)** | **Stratum-specific adjusted OR<sup>1</sup>** | **(95% CI)** | **Wald test P-value** | **Likelihood ratio test for interaction P-value** |
| --- | --- | --- | --- | --- | --- |
| *5 years and older* |  |  |  |  |  |
| No sore throat at onset | 21% (14/68) | 1.00 | . |  |  |
| **Sore throat at onset** | **42% (34/81)** | **2.79** | **(1.34-5.82)** | **0.006** | **0.009** |
| *Under 5 years old* |  |  |  |  |  |
| No Sore throat at onset | 31% (11/35) | 1.00 | . |  |  |
| **Sore throat at onset** | **17% (8/47)** | **0.65** | **(0.27-1.55)** | **0.328** |  |

**Supplementary Figure S1.**
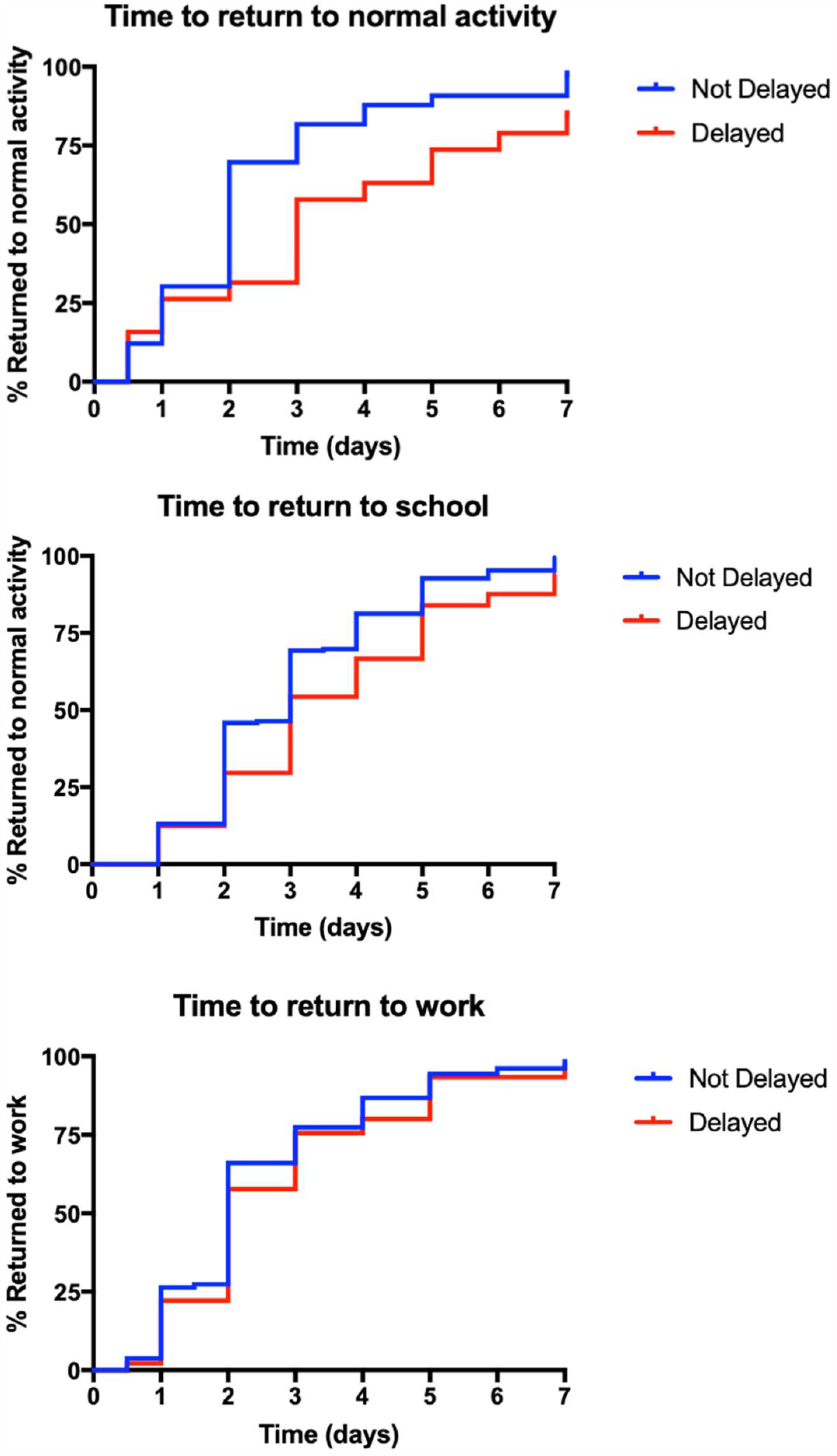
Recovery of scarlet fever cases among those in whom diagnosis was delayed or not delayed beyond first consultation with a health professional. Days elapsed from onset of symptoms to clinical recovery (n=52, Panel A), return to school (n=298, Panel B), and return to work for carers (n=161 Panel C).

### Footnotes

1 Public Health England, *Guidelines for the public health management of scarlet fever outbreaks in schools, nurseries and other childcare settings*, PHE, Editor. 2017, PHE: Wellington House, London.

## Notes

### Competing Interest Statement

The authors have declared no competing interest.

### Author Declarations

Ethical approval was provided by the London-Chelsea Research Ethics Committee, Reference 18/LO/0025; IRAS Reference 225006).

