## Supplementary File 1: Questionnaire 2018 for "Clinical management and impact of scarlet fever in the modern era: findings from a cross-sectional study of cases in London, 2018-2019"

Public Health England

### new scarlet fever

#### Introduction

Thank you for your interest in our research on scarlet fever. Scarlet fever is not normally a dangerous infection but it is quite infectious and spreads easily.

We would like to collect more information about each case of suspected scarlet fever so that we can describe the symptoms better, find out how quickly children respond to treatment, and measure the effect on families' everyday lives. This is important because the number of cases of scarlet fever has risen in the last few years and we need to be sure that the best advice is provided to families, schools and doctors.

The survey is entirely voluntary and your decision has no effect on the treatment that you and your child have in future.

It should take no longer than 15 minutes. Your child's name will not be put on the questionnaire. This means that no one can identify you or your child from this.

The survey is completely confidential; it is designed so that you cannot be identified even by the research team who have contacted you. If there are questions that you prefer not to answer then please just leave those out or tick the box 'prefer not to say'.

**Study Contacts**
Head of Operations, NIHR Health Protection Research Unit, Section of Infectious Diseases, Imperial College London, Du Cane Road, London W12 0NN.

Scarlet Fever Study Nurse, South London Health Protection Team, Public Health England, Zone C, 3rd Floor, Skipton House, 80 London Road, London SE1 6LH Telephone 0344 326 2052

Public Health England

new scarlet fever

#### Section A : About your child

1. **Enter your child's age at time of scarlet fever***

[ ]

2. **Gender**

○ Male ○ Female ○ Prefer not to say

3. **Ethnicity**

○ White British
○ White Irish
○ White: any other background
○ Mixed White and Black Caribbean
○ Mixed White and Black African
○ Mixed White and Asian
○ Mixed Any other mixed background
○ Asian or Asian British Indian
○ Asian or Asian British Pakistani
○ Asian or Asian British Bangladeshi
○ Asian/Asian British Any other Asian background
○ Black or Black British Caribbean
○ Black or Black British African
○ Black or Black British Any other Black background
○ Other Ethnic Group: Chinese
○ Any Other Ethnic Group
○ Prefer not to say

4. **At the time of having scarlet fever, was your child attending/did your child attend any of the following? (please tick one)**

☐ Playgroup/toddler group ☐ Day nursery ☐ School nursery
☐ Other type of nursery ☐ School Reception class

5. **If school nursery or reception, when did your child start school? (month/year)**

mm/yyyy

6. **If in school above Reception class, please state Year Group (1-13)**

### Your child's usual health (before scarlet fever suspected)

7. **Prior to scarlet fever, was your child normally quite healthy?**

○ Yes ○ No
○ Other, please specify

8. **If your child was already at school or nursery, approximately how many days off due to illness did they have in the 12 months BEFORE the suspected scarlet fever?**

9. **How many times did your child have a sore throat or tonsillitis in the 12 months BEFORE the suspected scarlet fever?**

○ 0 ○ 1 ○ 2 ○ 3 ○ 4

○ 5 ○ More than 5 ○ Can't say

10. **If your child has had more than one episode of sore throat or tonsillitis in the last 12 months, was your child diagnosed with group A streptococcus?**

○ Yes ○ No ○ Not sure

11. **Has your child had a tonsillectomy (tonsils removed)?**

○ Yes ○ No

12. **Has your child been admitted to hospital for any condition, other than this episode of scarlet fever, in the past?**

○ Yes ○ No ○ Prefer not to say

○ If yes, please tell us why...

13. **Was your child under follow up at a specialist clinic?**

○ Yes ○ No ○ Prefer not to say

○ If yes, please tell us why...

14. **Please tell us your relation to the child?**

○ Parent ○ Guardian ○ Temporary carer

○ Other, please specify

15. **How many other children under 16 are living with your child?**

○ 0 ○ 1 ○ 2 ○ 3 ○ 4

○ more than 4

Public Health England

**new scarlet fever**

**Section B About your child's illness (suspected scarlet fever)**

16. **Did your child have a rash?**

○ Yes

○ No

17. **If your child had a rash, where did the rash first appear?**

○ Face ○ Body ○ Arms/legs ○ Can't say

○ Other, please specify

18. **What was the rash like? Tick any that apply**

- ☐ red
- ☐ pink
- ☐ sand-papery/rough to feel
- ☐ small spots
- ☐ peeled off
- ☐ itchy
- ☐ Can't say
- ☐ Other, please specify

19. **How many days did the rash last in total? (give number of days, or indicate if you can't say)**

20. **If you child had an antibiotic how many days after starting the antibiotic did the rash go away? (give number of days or indicate if you can't say)**

21. **Did your child have a sore throat or difficulty swallowing?**

○ Yes ○ No ○ Can't say

22. **How many days did this last in total? (give number of days or indicate if you can't say)**

23. **Did your child have enlarged tonsils?**

○ Yes ○ No ○ Can't say

24. **Did your child have pus (white spots) on their tonsils?**

○ Yes ○ No ○ Can't say

25. **Was this the very first time your child has had a sore throat or tonsillitis?**

○ Yes ○ No ○ Not sure

26. **Did your child have a fever (a temperature above 37.5 C or above 99 F or shivering/feeling very hot to touch)?**

○ Yes ○ No ○ Not sure

27. **What was the highest temperature measured? (give measurement or indicate if you can't say)**

28. **How many days did the fever last in total? (give number of days or state if you can't say)**

29. **Did your child have an over the counter medication to help with fever (for example Calpol or Nurofen)?**

◯ Yes ◯ No ◯ Can't say

30. **How many days after starting the over-the-counter medicine did the fever go away? (give number of days or state if you can't say)**

31. **What other symptoms did your child have? Tick all that apply**

☐ Vomiting ☐ Diarrhoea ☐ Not eating ☐ Not playing ☐ Headache
☐ Tiredness ☐ Stomach ache ☐ Sore tongue ☐ Swollen tongue ☐ Earache
☐ Other, please describe

32. **What was the very first symptom you or your child noticed?**

☐ Rash ☐ Sore throat
☐ Fever ☐ Not playing/tiredness
☐ Can't say
☐ Other, please describe

33. **Was your child in contact with children with suspected scarlet fever before they fell ill?**

◯ Yes ◯ No ◯ Not sure

34. **Where was the contact?**

☐ nursery ☐ school
☐ neighbourhood ☐ family home
☐ Other, please specify

35. **Was your child in contact with other children with sore throat or tonsillitis before they fell ill?**

◯ Yes ◯ No ◯ Not sure

36. **Where was the contact?**

☐ nursery ☐ school
☐ neighbourhood ☐ family home
☐ Other, please specify

37. **Did your child have any other illness in the one month before they fell ill with suspected scarlet fever?**

◯ Yes ◯ No ◯ Not sure

38. **What did they have? Tick all that apply**

- ☐ common cold (runny nose)
- ☐ cough
- ☐ influenza (flu)
- ☐ vomiting or diarrhoea
- ☐ fever
- ☐ rash

Public Health England

**new scarlet fever**

**Section C: Use of healthcare for suspected scarlet fever**

39. **Where did you first seek advice when your child fell ill? Tick any that apply**

- ☐ GP/family doctor
- ☐ Local pharmacy
- ☐ School nurse
- ☐ Walk-in centre
- ☐ Urgent Care Centre
- ☐ Hospital Emergency Department
- ☐ NHS Direct
- ☐ Internet
- ☐ None of the above
- ☐ Other, please specify

40. **Did your child see a doctor or nurse for this illness?**

○ Yes ○ No

41. **With regard to the first time that your child saw a doctor or nurse, where were they based?**

- ☐ GP/usual GP practice
- ☐ Urgent Care Centre
- ☐ Emergency Department

42. **How many days after first symptom onset did your child first see a doctor/ nurse? (give number of days or state if you can't say)**

43. **Was a throat swab taken?**

- ☐ Yes
- ☐ No
- ☐ Can't say

44. **Did the swab show group A streptococcus..**

- ☐ yes
- ☐ no
- ☐ don't know
- ☐ Other result, please specify

45. **If your child had a complication or additional problem please tell us what it was...**

- ☐ throat/tonsil abscess
- ☐ otitis (ear infection)
- ☐ pneumonia (chest infection)
- ☐ meningitis
- ☐ septicaemia (blood infection)
- ☐ skin or soft tissue infection
- ☐ Other, please describe

46. **What was the suspected diagnosis when your child first saw a doctor? Tick any that apply**

☐ Sore throat ☐ Tonsillitis ☐ Influenza (flu)
☐ Measles ☐ A viral infection ☐ Scarlet fever
☐ Not sure
☐ Other, please specify

47. **Did your child need to see any doctor or nurse again for this illness?**

○ Yes ○ No

48. **Where were they based?**

☐ GP ☐ Urgent Care Centre ☐ Emergency Department
☐ Other, please specify

49. **Why did your child need to see a doctor again? Tick any that apply**

☐ Was asked to come back if no better ☐ Child developed a new symptom(s)
☐ Child could not take prescribed medicine ☐ Called back due to swab result
☐ Worried that it could be scarlet fever
☐ Other, please specify

50. **How many additional visits did you or your child make to any doctor related to suspected scarlet fever? (please tick one)**

○ 0 ○ 1 ○ 2 ○ 3 ○ 4 ○ 5 ○ 6 ○ 7 ○ 8 ○ 9 ○ 10
○ Can't say
○ Other, please specify

51. **Was your child admitted to hospital at any stage?**

○ Yes ○ No

52. **What was the reason for hospital admission?**

☐ Doctors unsure of diagnosis ☐ Fever was high ☐ Child developed a new symptom ☐ Child could not take prescribed medicine ☐ Child developed a complication
☐ Can't say
☐ Other, please specify

53. **How many nights did they stay in hospital? Give number of nights**

54. **Was your child admitted to an Intensive Care Unit or High Dependency Unit?**

○ Yes ○ No

55. **Was your child given antibiotics?**

○ Yes ○ No

56. **Were antibiotics given to start immediately, or antibiotics to collect if things did not improve (delayed antibiotics)?**

○ Yes immediate ○ Yes delayed ○ Can't say

57. **Please tell us the name of the antibiotic if possible:**

- ☐ Penicillin
- ☐ Amoxycillin
- ☐ Azithromycin
- ☐ Erythromycin
- ☐ Cephalexin
- ☐ Augmentin
- ☐ Can't say

58. **How many days was the antibiotic to be taken for?**

○ 1 ○ 2 ○ 3 ○ 4 ○ 5 ○ 6 ○ 7 ○ 8 ○ 9 ○ 10
○ Other, please specify

59. **How many times per day was it to be taken?**

○ 1 ○ 2 ○ 3 ○ 4 ○ Can't say
○ Other, please specify

60. **How many days did your child actually take the antibiotic for?**

○ 1 ○ 2 ○ 3 ○ 4 ○ 5 ○ 6 ○ 7 ○ 8 ○ 9 ○ 10
○ Other, please specify

61. **If your child did not take the antibiotic or took the antibiotic for fewer days than prescribed, please can you tell us why?**

- ☐ Was getting better anyway
- ☐ Difficult to collect prescription
- ☐ Stopped because my child got better
- ☐ Too many doses per day
- ☐ Not able to return to nursery/school on antibiotics
- ☐ Unpleasant taste
- ☐ Don't like giving my child medicines
- ☐ Can't say
- ☐ Other, please specify

62. **Did you use other medicines for child's symptoms? Tick all that apply**

- ☐ Paracetamol e.g. Calpol
- ☐ Ibuprofen e.g. Nurofen
- ☐ Nothing else
- ☐ Other, please specify

63. **If so how were these obtained?**

- ☐ Purchased by you
- ☐ Prescribed by your GP
- ☐ Other, please specify

64. **How many days did your child take these for?**

○ 1 ○ 2 ○ 3 ○ 4 ○ 5 ○ 6 ○ 7 ○ 8 ○ 9 ○ 10
○ Can't say
○ Other, please specify

Public Health
England

**new scarlet fever**

**Section D How your child's illness affected other people**

65. **Did your child miss days at nursery/school due to this illness?**
○ Yes ○ No

66. **How many days of school or nursery, that they would normally have attended, did your child miss?**
○ 1 ○ 2 ○ 3 ○ 4 ○ 5
○ 6 ○ 7 ○ More than 7 ○ Can't say
○ Other, please specify

67. **Did you miss time from work due to your child's illness?**
○ Yes ○ No

68. **If so, how many days did you miss?**
○ 1 ○ 2 ○ 3 ○ 4 ○ 5
○ 6 ○ 7 ○ More than 7 ○ Can't say
○ Other, please specify

69. **If so, was this to provide care for your sick child?**
○ Yes ○ No

70. **Did you become ill during your child's illness?**
○ Yes ○ No

71. **Did you lose income when away from worK?**
○ Yes ○ No

72. **Did you have to use any of the following to take time from work?**
☐ annual leave ☐ sick leave
☐ compassionate leave ☐ family leave
☐ Other, please specify

73. **If so, how many days did you use?**
○ 1 ○ 2 ○ 3 ○ 4 ○ 5
○ 6 ○ 7 ○ More than 7 ○ Can't say
○ Other, please specify

74. **Did another caregiver miss time from work due to your child's illness?**

○ Yes ○ No

75. **If so, was this to provide care for your sick child?**

○ Yes ○ No

76. **Did they become ill during your child's illness?**

○ Yes ○ No

77. **Did they lose income when away from work?**

○ Yes ○ No

78. **Did they use any of the following to take time off work?**

☐ annual leave ☐ sick leave
☐ compassionate leave ☐ family leave
☐ Other, please specify

79. **If so, how many days did they use?**

○ 1 ○ 2 ○ 3 ○ 4 ○ 5
○ 6 ○ 7 ○ More than 7 ○ Can't say
○ Other, please specify

80. **Did you have to find alternative childcare due to your child's illness?**

○ Yes ○ No

81. **If so, who was this?**

☐ Friend ☐ Family member ☐ Professional (paid for) childcare
☐ Other, please specify

82. **If so, how many days did you need help?**

○ 1 ○ 2 ○ 3 ○ 4 ○ 5
○ 6 ○ 7 ○ More than 7 ○ Can't say
○ Other, please specify

83. **Did any of your other children miss days at nursery/school due to your child's illness?**

○ Yes ○ No

84. **If so, how many days did other children miss?**

○ 1 ○ 2 ○ 3 ○ 4 ○ 5
○ 6 ○ 7 ○ More than 7 ○ Can't say
○ Other, please specify

85. **If so, why did this happen?**

☐ Couldn't leave my sick child alone to take others to school/nursery
☐ Worried that my other children were also sick

☐ Older children helped to look after younger child
☐ Other, please specify

86. **We are interested to know how a diagnosis of suspected scarlet fever affected how you felt.**

**Had you heard of scarlet fever before?**

○ Yes ○ No

87. **If yes, what was your impression about it? Tick all that apply**

☐ Historical and dangerous illness
☐ Historical illness that disappeared a century ago
☐ Historical illness that is making a comeback
☐ Historical illness that never went away
☐ A childhood illness with rash that is rare
☐ A childhood illness with rash that can be very infectious
☐ A childhood illness that can be treated with antibiotics
☐ A childhood illness that cannot at present be prevented by vaccination
☐ A childhood illness caused by streptococcal bacteria
☐ Anything else?

88. **Had you heard of group A streptococcus before?**

○ Yes ○ No

89. **How worried were you on a scale of 0 - 10? (where 0 is not worried at all and 10 is the most worried you could ever be about your child)**

0

90. **If yes, what was your impression about it? Tick all that apply.**

☐ Can cause sore throat & tonsillitis
☐ Causes fewer sore throats than viruses
☐ Can cause impetigo (skin condition)
☐ Can infect wounds
☐ Can rarely cause more dangerous infections
☐ Rare serious infections include necrotising fasciitis and toxic shock
☐ Can be treated with penicillin
☐ There is currently no vaccine against it
☐ Anything else?

91. **How did other people react to your child's illness? Tick all that apply**

☐ No reaction
☐ Less worried than me
☐ More worried than me
☐ Contacted me to ask how my child was
☐ Offered to help
☐ Kept their child away from my child
☐ Other, please specify

92. **What was the most helpful source of information for you? Tick all that apply**

☐ GP/family doctor
☐ Local pharmacy
☐ School/nursery
☐ Urgent Care Centre
☐ Hospital Emergency Department
☐ Family/friends
☐ NHS Direct
☐ Internet
☐ Public Health England
☐ None of these
☐ Other, please specify

93. **Is there anything else that you would like to tell us about your child's illness or the impact it had on your or your child?**

**Thank you so much for your help in completing the survey. This will help us to improve services and preventative strategies in the future when dealing with cases of scarlet fever. Please click done to submit your answers.**
