## Supplementary File 2: Questionnaire 2019 for "Clinical management and impact of scarlet fever in the modern era: findings from a cross-sectional study of cases in London, 2018-2019"

Public Health England

### School Survey of Scarlet fever

#### Introduction

Thank you for your interest in our research on scarlet fever. Scarlet fever is not normally a dangerous infection however it is quite infectious and spreads easily.

We would like to collect more information about each child with suspected scarlet fever and on those children in the school who remain well. This will then allow us to describe the symptoms better, find out how quickly children respond to treatment, and measure the effect of scarlet fever infection on the everyday lives of families compared to other infections circulating at this time of the year. This is important because the number of cases of scarlet fever has risen in the last few years and we need to be sure that the best advice is provided to families, schools and doctors.

The survey is entirely voluntary, and your decision has no effect on the treatment that you and your child have in future.

Please note the questionnaire has been designed so that in some instances, depending on your response to specific questions you may miss questions and be directed to questions further on in the questionnaire. Additionally, should you not be able to answer any question, please do leave it blank and move to the next question.

It should take no longer than 15 minutes. Your child's name will not be put on the questionnaire and they will remain anonymous. This means that no one can identify you or your child from this.

The survey is completely confidential; it is designed so that you cannot be identified even by the research team who have contacted you. If there are questions that you prefer not to answer then please just leave those out or tick the box 'prefer not to say'.

**Study Contacts**

Head of Operations, NIHR Health Protection Research Unit, Section of Infectious Diseases, Imperial College London, Du Cane Road, London W12 0NN.

Scarlet Fever Study Nurse, South London Health Protection Team, Public Health England, Zone C, 3rd Floor, Skipton House, 80 London Road, London SE1 6LH Telephone 0344 326 2052

Public Health England

School Survey of Scarlet fever

### Section A : About your child

1. **How old is your child?***

(If you have more than one child at this school/ nursery, please complete this survey separately for each child)

The value must be between 0 and 15, inclusive.

2. **Gender**

○ Male ○ Female ○ Prefer not to say

3. **Ethnicity**

○ White British
○ White Irish
○ White: any other background
○ Mixed White and Black Caribbean
○ Mixed White and Black African
○ Mixed White and Asian
○ Mixed Any other mixed background
○ Asian or Asian British Indian
○ Asian or Asian British Pakistani
○ Asian or Asian British Bangladeshi
○ Asian/Asian British Any other Asian background
○ Black or Black British Caribbean
○ Black or Black British African
○ Black or Black British Any other Black background
○ Other Ethnic Group: Chinese
○ Any Other Ethnic Group
○ Prefer not to say

4. **Does your child attend any of the following? (please tick all that apply)**

☐ Playgroup/toddler group
☐ School nursery
☐ Primary school Reception class
☐ Day nursery
☐ Other type of nursery
☐ Primary school Year 1-6

5. **How long has your child been at nursery or school in the UK?**

○ Less than 1 year
○ 1 - 2 years
○ 3 - 4 years
○ 5 years and over

6. **If your child is in a school class, please state the Year Group**

○ Infant
○ Reception
○ 1
○ 2
○ 3
○ 4
○ 5
○ 6

### Your child's usual health

7. **Is your child normally quite healthy?**

○ Yes ○ No

8. **Is your child under follow up at a specialist clinic?**
   - ○ Yes
   - ○ No
   - ○ Prefer not to say

9. **Has your child been admitted to hospital for any condition in the past?**
   - ○ Yes
   - ○ No
   - ○ Prefer not to say

10. **Approximately how many days off school or nursery due to illness has your child had in the last 12 months overall ?**
    - ○ Up to 2 days
    - ○ 3 - 6 days
    - ○ 1 - 2 weeks
    - ○ 3 - 4 weeks
    - ○ 5 weeks and over

11. **How many times has your child had a sore throat or tonsillitis in the last 12 months (including if they have one now)?**
    - ○ 0
    - ○ 1
    - ○ 2
    - ○ 3
    - ○ 4
    - ○ 5
    - ○ More than 5
    - ○ Can't say

12. **If your child has had more than one episode of sore throat or tonsillitis in the last 12 months, was your child diagnosed with group A streptococcus?**
    - ○ Yes
    - ○ No
    - ○ Not sure

13. **Has your child had a tonsillectomy (tonsils removed)?**
    - ○ Yes
    - ○ No

14. **As far as you know did your child receive a nasal flu vaccine this season (any time since September 2018)?**
    - ○ Yes
    - ○ No

15. **When did your child receive the nasal flu vaccine?**
    - ○ September 2018
    - ○ October 2018
    - ○ November 2018
    - ○ December 2018
    - ○ January 2019
    - ○ February 2019
    - ○ Don't know

16. **Please tell us your relationship to the child?**
    - ○ Parent
    - ○ Guardian
    - ○ Temporary carer
    - ○ Other, please specify

    [ ]

17. **How many people (including you) are living in the same household with your child?**
    - ○ 1
    - ○ 2
    - ○ 3
    - ○ 4
    - ○ more than 4

18.

**How many other children aged 11 and under are living in the same household with your child?**

○ 0 ○ 1 ○ 2 ○ 3 ○ 4
○ more than 4

Public Health England

### School Survey of Scarlet fever

**Section B About your child and your child's recent illness (suspected scarlet fever), if relevant**

19. **Has your child been ill in the last four weeks with fever, rash, sore throat or tonsillitis?**
○ Yes ○ No

20. **Has your child been diagnosed with scarlet fever by a doctor or nurse in the last four weeks?**
○ Yes
○ No

21. **Has your child had a new rash in the last four weeks? (not including long term skin conditions e.g. eczema)**
○ Yes
○ No

22. **If your child had a rash, where did the rash FIRST appear?**
○ Face ○ Body ○ Arms/legs ○ Don't know

23. **What was the rash like? Tick any that apply**
☐ red
☐ sand-papery/rough to feel
☐ peeled off
☐ Can't say
☐ Other, please specify
☐ pink
☐ small spots
☐ itchy

24. **How many days did the rash last in total? (give number of days)**

25. **As far as you know, has your child had a sore throat or difficulty swallowing in the last four weeks?**
○ Yes ○ No

26. **How many days did the sore throat or difficulty swallowing last in total? (give number of days)**

27. **As far as you know, has your child had enlarged tonsils in the last 4 weeks?**
- ○ Yes
- ○ No
- ○ Tonsils previously removed

28. **As far as you know, did your child have pus (white spots) on their tonsils?**
- ○ Yes
- ○ No
- ○ Tonsils previously removed

29. **As far as you know, has your child had a fever (a temperature above 37.5 C or shivering/feeling very hot to touch) in the last four weeks?**
- ○ Yes
- ○ No

30. **What was the highest temperature measured in Celsius? (give measurement or indicate if you don't know)**
- ○ Under 37.5 °C
- ○ 37.5 - 38 °C
- ○ 38.1 - 38.5 °C
- ○ 38.6 - 39 °C
- ○ 39.1 - 39.5 °C
- ○ More than 39.5 °C
- ○ don't know

31. **How many days did the fever last in total? (give number of days)**

[ ]

32. **As far as you know, did your child have an over the counter medication to help with fever (for example Calpol or Nurofen)?**
- ○ Yes
- ○ No

33. **As far as you know, has your child had impetigo (a common mild skin infection) in the last four weeks?**
- ○ Yes
- ○ No

34. **As far as you know, has your child had a flu like illness in the last four weeks?**
- ○ Yes
- ○ No

35. **As far as you know, has your child been in contact with anyone with suspected scarlet fever?**
- ○ Yes
- ○ No

36. **Where was the contact with scarlet fever?**

Tick all that apply
- ☐ nursery
- ☐ school
- ☐ neighbourhood
- ☐ family home
- ☐ Other, please specify

[ ]

37. **As far as you know, has your child been in contact with other children or adults with sore throat or tonsillitis in the last four weeks?**
- ○ Yes
- ○ No

38. **Where was the contact(s)?**

Tick all that apply

- ☐ nursery
- ☐ school
- ☐ neighbourhood
- ☐ family home
- ☐ Other, please specify

Public Health England

### School Survey of Scarlet fever

#### Section C: Use of healthcare for suspected scarlet fever

39. **When your child was ill in the last four weeks, where did you first seek advice ?**

Tick all that apply

- ☐ GP/family doctor
- ☐ Practice nurse
- ☐ Local pharmacy
- ☐ School nurse
- ☐ Walk-in centre
- ☐ Urgent Care Centre
- ☐ Hospital Emergency Department
- ☐ NHS Direct
- ☐ Internet
- ☐ None of the above
- ☐ Other, please specify

40. **How many days after first symptom onset did your child first see a doctor/ nurse? (give number of days)**

41. **As far as you know, was a throat swab taken?**

- ○ Yes
- ○ No

42. **Did the swab show group A streptococcus (the bacteria which causes Scarlet fever)?**

- ○ Yes
- ○ No
- ○ Don't know
- ○ Other result, please specify

43. **What was the suspected diagnosis when your child FIRST saw a doctor? Tick any that apply**

- ☐ Tonsillitis
- ☐ Influenza (flu)
- ☐ Measles
- ☐ A viral infection
- ☐ Scarlet fever
- ☐ Not sure
- ☐ Other, please specify

44. **Did your child need to see a doctor or nurse again for this illness?**

○ Yes ○ No

45. **Where were they based?**

- ☐ GP
- ☐ Urgent Care Centre
- ☐ Emergency Department

☐ Other, please specify

46. **Why did your child need to see a doctor again? Tick any that apply**

☐ Was asked to come back if not better
☐ Child developed a new symptom(s)
☐ Child could not take prescribed medicine
☐ Called back due to swab result
☐ Worried that it could be scarlet fever
☐ Other, please specify

47. **Has your child been admitted to hospital in the last four weeks?**

○ Yes ○ No

48. **What was the reason for hospital admission?**

Tick all that apply

☐ Doctors unsure of diagnosis
☐ Fever was high
☐ Child developed a new symptom
☐ Child could not take prescribed medicine
☐ Child developed a complication
☐ Other, please specify

49. **How many nights did they stay in hospital? Give number of nights**

50. **Was your child given antibiotics?**

○ Yes
○ No
○ Don't know

51. **When were antibiotics taken?**

○ Just in hospital
○ In hospital and to take home
○ Just to take at home

52. **Were you advised to start antibiotics immediately or to start only if things did not improve?**

○ Yes immediate ○ Yes delayed ○ Can't say

53. **How many days after the first symptom onset did your child START to take antibiotics (give number of days )**

54. **How many days after rash onset did your child START TO take antibiotics (give number of days)**

State number of days or if child did not have a rash

○ 0-1
○ 2
○ 3
○ 4
○ 5
○ 6 or more
○ Child did not have a rash

55. **How many days was your child unwell for AFTER they had started their antibiotics?**

- ○ 0-1 days
- ○ 2
- ○ 3
- ○ 4
- ○ 5
- ○ 6 or more
- ○ Not applicable

56. **Please tell us the name of the antibiotic if possible:**
- ○ Penicillin
- ○ Amoxycillin
- ○ Azithromycin
- ○ Erythromycin
- ○ Cephalexin
- ○ Augmentin
- ○ Can't say
- ○ Other, please specify

57. **Please tell us the dose of your antibiotic if possible:**
- ○ 125mg
- ○ 250mg
- ○ 375mg
- ○ 500mg
- ○ 1 g
- ○ Don't know

58. **How many times per day was it to be taken?**
○ 1 ○ 2 ○ 3 ○ 4 ○ Can't say

59. **How many days was the antibiotic to be taken for?**
○ 1 ○ 2 ○ 3 ○ 4 ○ 5 ○ 6 ○ 7 ○ 8 ○ 9 ○ 10

60. **How many days did your child ACTUALLY take the antibiotic for?**
○ 1 ○ 2 ○ 3 ○ 4 ○ 5 ○ 6 ○ 7 ○ 8 ○ 9 ○ 10

61. **If your child did not take the antibiotic or took the antibiotic for fewer days than prescribed, please can you tell us why?**
Tick all that apply
- ☐ Was getting better anyway
- ☐ Stopped because my child got better
- ☐ Not able to return to nursery/school on antibiotics
- ☐ Don't like giving my child medicines
- ☐ Other, please specify
- ☐ Difficult to collect prescription
- ☐ Too many doses per day
- ☐ Unpleasant taste

62. **Did you use other medicines for child's symptoms? Tick all that apply**
- ☐ Paracetamol e.g. Calpol
- ☐ Ibuprofen e.g. Nurofen
- ☐ Nothing else
- ☐ Other, please specify

Public Health England

### School Survey of Scarlet fever

#### Section D How your child's illness affected other people

63. **If your child was ill, did your child miss days at nursery/school?**
   - ○ Yes
   - ○ No
   - ○ Not applicable

64. **How many days of school or nursery, that they would normally have attended, did your child miss?**
   - ○ 1
   - ○ 2
   - ○ 3
   - ○ 4
   - ○ 5
   - ○ 6
   - ○ 7
   - ○ More than 7
   - ○ Not applicable, child not ill

65. **How many days after starting antibiotics did your child feel well enough to resume normal activities, such as going to school or nursery?**
   - ○ 0
   - ○ 1
   - ○ 2
   - ○ 3
   - ○ 4
   - ○ 5
   - ○ 6
   - ○ 7
   - ○ More than 7
   - ○ Not applicable, my child was not ill

66. **Did you become ill during your child's illness?**
   - ○ Yes
   - ○ No
   - ○ Not applicable

67. **Did you miss time from work to provide care for your sick child?**
   - ○ Yes
   - ○ No
   - ○ Not applicable

68. **If so, how many days did you miss?**
   - ○ 1
   - ○ 2
   - ○ 3
   - ○ 4
   - ○ 5
   - ○ 6
   - ○ 7
   - ○ More than 7

69. **As a result, did you lose income when away from work?**
   - ☐ Yes
   - ☐ No
   - ☐ Not applicable

70. **Did you have to use any of the following to take time from work to care for your sick child?**

- [ ] annual leave
- [ ] sick leave
- [ ] compassionate leave
- [ ] family leave
- [ ] Other, please specify

71. **If so, how many days did you use?**
○ 1 ○ 2 ○ 3 ○ 4 ○ 5
○ 6 ○ 7 ○ More than 7

72. **Did another caregiver miss time from work to care for your sick child?**
○ Yes ○ No

73. **As a result, did they lose income when away from work?**
○ Yes ○ No

74. **Did they use any of the following to take time off work to care for your sick child?**
- [ ] annual leave
- [ ] sick leave
- [ ] compassionate leave
- [ ] family leave
- [ ] Other, please specify

75. **If so, how many days did they use?**
○ 1 ○ 2 ○ 3 ○ 4 ○ 5
○ 6 ○ 7 ○ More than 7 ○ Can't say

76. **Did they become ill during your child's illness?**
○ Yes ○ No

77. **Did you have to find alternative childcare due to your child's illness?**
○ Yes ○ No

78. **If so, who was this?**
○ Friend
○ Family member
○ Professional (paid for) childcare
○ Other, please specify

79. **If so, how many days did you need help?**
○ 1 ○ 2 ○ 3 ○ 4 ○ 5
○ 6 ○ 7 ○ More than 7

80. **Did any of your other children miss days at nursery/school due to your child's illness?**
○ Yes ○ No

81. **If so, how many days did other children miss?**
○ 1 ○ 2 ○ 3 ○ 4 ○ 5
○ 6 ○ 7 ○ More than 7

82. **If so, why did this happen?**
- [ ] Couldn't leave my sick child alone to take others to school/nursery

☐ Worried that my other children were also sick
☐ Older children helped to look after younger child
☐ Other, please specify

83. **Before you had this letter from the school or nursery, had you heard of scarlet fever before?**
○ Yes ○ No

84. **If yes, what was your impression about it? Tick all that apply**
☐ Historical and dangerous illness
☐ Historical illness that is making a comeback
☐ A childhood illness with rash that is rare
☐ A childhood illness that can be treated with antibiotics
☐ A childhood illness caused by streptococcal bacteria
☐ Historical illness that disappeared a century ago
☐ Historical illness that never went away
☐ A childhood illness with rash that can be very infectious
☐ A childhood illness that cannot at present be prevented by vaccination
☐ Anything else?

85. **If yes, what was your impression about it? Tick all that apply.**
☐ Can cause sore throat & tonsillitis
☐ Can cause impetigo (skin condition)
☐ Can rarely cause more dangerous infections
☐ Can be treated with penicillin
☐ Causes fewer sore throats than viruses
☐ Can infect wounds
☐ Rare serious infections include necrotising fasciitis and toxic shock
☐ There is currently no vaccine against it
☐ Anything else?

86. **What are the most helpful sources of information for you when your child is unwell? Tick all that apply**
☐ GP/family doctor
☐ Urgent Care Centre
☐ NHS Direct
☐ Local pharmacy
☐ Hospital Emergency Department
☐ Internet
☐ School/nursery
☐ Family/friends
☐ Public Health England
☐ None of these
☐ Other, please specify

87. **Do you now think your child may have had scarlet fever in the last four weeks?**
○ Yes ○ No

88. **Do you think anyone else in your family or household has had any of the following in the last four weeks?**
☐ Sore throat
☐ Tonsillitis
☐ Impetigo
☐ Scarlet Fever
☐ Cellulitis
☐ Other, please specify

89. **If your child was unwell, is there anything else that you would like to tell us about your child's illness or the impact it had on your or your child?**

**Thank you so much for your help in completing the survey. This will help us to improve services and preventative strategies in the future when dealing with cases of scarlet fever. Please click done to submit your answers.**
