## Supplementary material for "Clinical management and impact of scarlet fever in the modern era: findings from a cross-sectional study of cases in London, 2018-2019": Disclosure Forms

#### ICMJE DISCLOSURE FORM

**Date:_24/6/2021______________________________________________________________________**
**Your Name: M. Trent Herdman_______________________________________________________________________**
**Manuscript Title: Clinical management and impact of scarlet fever in the modern era: findings from a cross-sectional study of cases in London, 2018-2019.**
**Manuscript number (if known):_NA__________________________________________________________________**

**In the interest of transparency, we ask you to disclose all relationships/activities/interests listed below that are related to the content of your manuscript. "Related" means any relation with for-profit or not-for-profit third parties whose interests may be affected by the content of the manuscript. Disclosure represents a commitment to transparency and does not necessarily indicate a bias. If you are in doubt about whether to list a relationship/activity/interest, it is preferable that you do so.**

**The following questions apply to the author's relationships/activities/interests as they relate to the current manuscript only.**

**The author's relationships/activities/interests should be defined broadly. For example, if your manuscript pertains to the epidemiology of hypertension, you should declare all relationships with manufacturers of antihypertensive medication, even if that medication is not mentioned in the manuscript.**

**In item #1 below, report all support for the work reported in this manuscript without time limit. For all other items, the time frame for disclosure is the past 36 months.**

| | | **Name all entities with whom you have this relationship or indicate none (add rows as needed)** | **Specifications/Comments (e.g., if payments were made to you or to your institution)** |
|---|---|---|---|
| | | **Time frame: Since the initial planning of the work** | |
| 1 | All support for the present manuscript (e.g., funding, provision of study materials, medical writing, article processing charges, etc.) **No time limit for this item.** | ___X_None | |
| | | **Time frame: past 36 months** | |
| 2 | Grants or contracts from any entity (if not indicated in item #1 above). | __X__None | |
| 3 | Royalties or licenses | __X__None | |
| 4 | Consulting fees | __X__None | |

| | | |
|---|---|---|
| 5 | Payment or honoraria for lectures, presentations, speakers bureaus, manuscript writing or educational events | __X__None |
| 6 | Payment for expert testimony | __X__None |
| 7 | Support for attending meetings and/or travel | __X__None |
| 8 | Patents planned, issued or pending | __X__None |
| 9 | Participation on a Data Safety Monitoring Board or Advisory Board | __X__None |
| 10 | Leadership or fiduciary role in other board, society, committee or advocacy group, paid or unpaid | __X__None |
| 11 | Stock or stock options | _X___None |
| 12 | Receipt of equipment, materials, drugs, medical writing, gifts or other services | _X___None |
| 13 | Other financial or non-financial interests | __X__None |

**Please place an "X" next to the following statement to indicate your agreement:**

**__X_ I certify that I have answered every question and have not altered the wording of any of the questions on this form.**

#### ICMJE DISCLOSURE FORM

**Date:_24/6/2021___________________________________________________________________________**
**Your Name: Rebecca Cordery____________________________________________________________________**
**Manuscript Title: Clinical management and impact of scarlet fever in the modern era: findings from a cross-sectional study of cases in London, 2018-2019.**
**Manuscript number (if known):_NA_______________________________________________________________**

**In the interest of transparency, we ask you to disclose all relationships/activities/interests listed below that are related to the content of your manuscript. "Related" means any relation with for-profit or not-for-profit third parties whose interests may be affected by the content of the manuscript. Disclosure represents a commitment to transparency and does not necessarily indicate a bias. If you are in doubt about whether to list a relationship/activity/interest, it is preferable that you do so.**

**The following questions apply to the author's relationships/activities/interests as they relate to the current manuscript only.**

**The author's relationships/activities/interests should be defined broadly. For example, if your manuscript pertains to the epidemiology of hypertension, you should declare all relationships with manufacturers of antihypertensive medication, even if that medication is not mentioned in the manuscript.**

**In item #1 below, report all support for the work reported in this manuscript without time limit. For all other items, the time frame for disclosure is the past 36 months.**

| | | **Name all entities with whom you have this relationship or indicate none (add rows as needed)** | **Specifications/Comments (e.g., if payments were made to you or to your institution)** |
|---|---|---|---|
| | | **Time frame: Since the initial planning of the work** | |
| 1 | All support for the present manuscript (e.g., funding, provision of study materials, medical writing, article processing charges, etc.) **No time limit for this item.** | ___X_None | |
| | | **Time frame: past 36 months** | |
| 2 | Grants or contracts from any entity (if not indicated in item #1 above). | __X__None | |
| 3 | Royalties or licenses | __X__None | |
| 4 | Consulting fees | __X__None | |

| | | |
|---|---|---|
| 5 | Payment or honoraria for lectures, presentations, speakers bureaus, manuscript writing or educational events | __X__None |
| 6 | Payment for expert testimony | __X__None |
| 7 | Support for attending meetings and/or travel | __X__None |
| 8 | Patents planned, issued or pending | __X__None |
| 9 | Participation on a Data Safety Monitoring Board or Advisory Board | __X__None |
| 10 | Leadership or fiduciary role in other board, society, committee or advocacy group, paid or unpaid | __X__None |
| 11 | Stock or stock options | _X___None |
| 12 | Receipt of equipment, materials, drugs, medical writing, gifts or other services | _X___None |
| 13 | Other financial or non-financial interests | __X__None |

**Please place an "X" next to the following statement to indicate your agreement:**

**__X_ I certify that I have answered every question and have not altered the wording of any of the questions on this form.**

#### ICMJE DISCLOSURE FORM

**Date:17 June 2021**______________________________________________________________
**Your Name: Basel Karo**______________________________________________________
**Manuscript Title: Clinical management and impact of scarlet fever in the modern era: findings from a cross-sectional study of cases in London, 2018-2019.**
**Manuscript number (if known):NA** ____________________________________________________

**In the interest of transparency, we ask you to disclose all relationships/activities/interests listed below that are related to the content of your manuscript. "Related" means any relation with for-profit or not-for-profit third parties whose interests may be affected by the content of the manuscript. Disclosure represents a commitment to transparency and does not necessarily indicate a bias. If you are in doubt about whether to list a relationship/activity/interest, it is preferable that you do so.**

**The following questions apply to the author's relationships/activities/interests as they relate to the current manuscript only.**

**The author's relationships/activities/interests should be defined broadly. For example, if your manuscript pertains to the epidemiology of hypertension, you should declare all relationships with manufacturers of antihypertensive medication, even if that medication is not mentioned in the manuscript.**

**In item #1 below, report all support for the work reported in this manuscript without time limit. For all other items, the time frame for disclosure is the past 36 months.**

| | | **Name all entities with whom you have this relationship or indicate none (add rows as needed)** | **Specifications/Comments (e.g., if payments were made to you or to your institution)** |
|---|---|---|---|
| | | **Time frame: Since the initial planning of the work** | |
| 1 | All support for the present manuscript (e.g., funding, provision of study materials, medical writing, article processing charges, etc.) **No time limit for this item.** | ____ None | |
| | | **Time frame: past 36 months** | |
| 2 | Grants or contracts from any entity (if not indicated in item #1 above). | ____ None | |
| 3 | Royalties or licenses | ____ None | |

| | | |
|---|---|---|
| 4 | Consulting fees | ____ None |
| 5 | Payment or honoraria for lectures, presentations, speakers bureaus, manuscript writing or educational events | ____ None |
| 6 | Payment for expert testimony | ____ None |
| 7 | Support for attending meetings and/or travel | ____ None |
| 8 | Patents planned, issued or pending | ____ None |
| 9 | Participation on a Data Safety Monitoring Board or Advisory Board | ____ None |
| 10 | Leadership or fiduciary role in other board, society, committee or advocacy group, paid or unpaid | ____ None |
| 11 | Stock or stock options | ____ None |
| 12 | Receipt of equipment, materials, drugs, medical writing, gifts or other services | ____ None |
| 13 | Other financial or non-financial interests | ____ None |

**Please place an "X" next to the following statement to indicate your agreement:**

**_X_ I certify that I have answered every question and have not altered the wording of any of the questions on this form.**

#### ICMJE DISCLOSURE FORM

**Date:17 June 2021__________________________________________________________________________**
**Your Name:_Ms Amrit Kaur Purba**
**Manuscript Title: Clinical management and impact of scarlet fever in the modern era: findings from a cross-sectional study of cases in London, 2018-2019.**
**Manuscript number (if known):NA ________________________________________________________________**

**In the interest of transparency, we ask you to disclose all relationships/activities/interests listed below that are related to the content of your manuscript. "Related" means any relation with for-profit or not-for-profit third parties whose interests may be affected by the content of the manuscript. Disclosure represents a commitment to transparency and does not necessarily indicate a bias. If you are in doubt about whether to list a relationship/activity/interest, it is preferable that you do so.**

**The following questions apply to the author's relationships/activities/interests as they relate to the current manuscript only.**

**The author's relationships/activities/interests should be defined broadly. For example, if your manuscript pertains to the epidemiology of hypertension, you should declare all relationships with manufacturers of antihypertensive medication, even if that medication is not mentioned in the manuscript.**

**In item #1 below, report all support for the work reported in this manuscript without time limit. For all other items, the time frame for disclosure is the past 36 months.**

| | | **Name all entities with whom you have this relationship or indicate none (add rows as needed)** | **Specifications/Comments (e.g., if payments were made to you or to your institution)** |
|---|---|---|---|
| | | **Time frame: Since the initial planning of the work** | |
| 1 | All support for the present manuscript (e.g., funding, provision of study materials, medical writing, article processing charges, etc.) **No time limit for this item.** | Medical Research Council | MC_UU_00022/2 |
| | | **Time frame: past 36 months** | |
| 2 | Grants or contracts from any entity (if not indicated in item #1 above). | See above | |
| 3 | Royalties or licenses | ____ None | |

| | | |
|---|---|---|
| 4 | Consulting fees | ____ None |
| 5 | Payment or honoraria for lectures, presentations, speakers bureaus, manuscript writing or educational events | ____ None |
| 6 | Payment for expert testimony | ____ None |
| 7 | Support for attending meetings and/or travel | ____ None |
| 8 | Patents planned, issued or pending | ____ None |
| 9 | Participation on a Data Safety Monitoring Board or Advisory Board | ____ None |
| 10 | Leadership or fiduciary role in other board, society, committee or advocacy group, paid or unpaid | ____ None |
| 11 | Stock or stock options | ____ None |
| 12 | Receipt of equipment, materials, drugs, medical writing, gifts or other services | ____ None |
| 13 | Other financial or non-financial interests | ____ None |

**Please place an "X" next to the following statement to indicate your agreement:**

**__X_ I certify that I have answered every question and have not altered the wording of any of the questions on this form.**

### ICMJE DISCLOSURE FORM

**Date: 21st June 2021**____________________________________________________________
**Your Name:__Lipi Begum**_____________________________________________________
**Manuscript Title: Clinical management and impact of scarlet fever in the modern era: findings from a cross-sectional study of cases in London, 2018-2019.**
**Manuscript number (if known):NA** __________________________________________________

**In the interest of transparency, we ask you to disclose all relationships/activities/interests listed below that are related to the content of your manuscript. "Related" means any relation with for-profit or not-for-profit third parties whose interests may be affected by the content of the manuscript. Disclosure represents a commitment to transparency and does not necessarily indicate a bias. If you are in doubt about whether to list a relationship/activity/interest, it is preferable that you do so.**

**The following questions apply to the author's relationships/activities/interests as they relate to the current manuscript only.**

**The author's relationships/activities/interests should be defined broadly. For example, if your manuscript pertains to the epidemiology of hypertension, you should declare all relationships with manufacturers of antihypertensive medication, even if that medication is not mentioned in the manuscript.**

**In item #1 below, report all support for the work reported in this manuscript without time limit. For all other items, the time frame for disclosure is the past 36 months.**

| | | **Name all entities with whom you have this relationship or indicate none (add rows as needed)** | **Specifications/Comments (e.g., if payments were made to you or to your institution)** |
|---|---|---|---|
| **Time frame: Since the initial planning of the work** | | | |
| 1 | All support for the present manuscript (e.g., funding, provision of study materials, medical writing, article processing charges, etc.) **No time limit for this item.** | ____ None | |
| **Time frame: past 36 months** | | | |
| 2 | Grants or contracts from any entity (if not indicated in item #1 above). | ____ None | |
| 3 | Royalties or licenses | ____ None | |

| | | |
|---|---|---|
| 4 | Consulting fees | ____ None |
| 5 | Payment or honoraria for lectures, presentations, speakers bureaus, manuscript writing or educational events | ____ None |
| 6 | Payment for expert testimony | ____ None |
| 7 | Support for attending meetings and/or travel | ____ None |
| 8 | Patents planned, issued or pending | ____ None |
| 9 | Participation on a Data Safety Monitoring Board or Advisory Board | ____ None |
| 10 | Leadership or fiduciary role in other board, society, committee or advocacy group, paid or unpaid | ____ None |
| 11 | Stock or stock options | ____ None |
| 12 | Receipt of equipment, materials, drugs, medical writing, gifts or other services | ____ None |
| 13 | Other financial or non-financial interests | ____ None |

**Please place an "X" next to the following statement to indicate your agreement:**

**__X_ I certify that I have answered every question and have not altered the wording of any of the questions on this form.**

#### ICMJE DISCLOSURE FORM

**Date: 21 June 2021____________________________________________________________________**
**Your Name:___Theresa Lamagni __________________________________________________________**
**Manuscript Title: Clinical management and impact of scarlet fever in the modern era: findings from a cross-sectional study of cases in London, 2018-2019.**
**Manuscript number (if known):NA __________________________________________________________**

**In the interest of transparency, we ask you to disclose all relationships/activities/interests listed below that are related to the content of your manuscript. "Related" means any relation with for-profit or not-for-profit third parties whose interests may be affected by the content of the manuscript. Disclosure represents a commitment to transparency and does not necessarily indicate a bias. If you are in doubt about whether to list a relationship/activity/interest, it is preferable that you do so.**

**The following questions apply to the author's relationships/activities/interests as they relate to the current manuscript only.**

**The author's relationships/activities/interests should be defined broadly. For example, if your manuscript pertains to the epidemiology of hypertension, you should declare all relationships with manufacturers of antihypertensive medication, even if that medication is not mentioned in the manuscript.**

**In item #1 below, report all support for the work reported in this manuscript without time limit. For all other items, the time frame for disclosure is the past 36 months.**

| | | Name all entities with whom you have this relationship or indicate none (add rows as needed) | Specifications/Comments (e.g., if payments were made to you or to your institution) |
|---|---|---|---|
| | | **Time frame: Since the initial planning of the work** | |
| 1 | All support for the present manuscript (e.g., funding, provision of study materials, medical writing, article processing charges, etc.) **No time limit for this item.** | _x___ None | |
| | | **Time frame: past 36 months** | |
| 2 | Grants or contracts from any entity (if not indicated in item #1 above). | __x__ None | |
| 3 | Royalties or licenses | __x__ None | |

| | | |
|---|---|---|
| 4 | Consulting fees | __x__ None |
| 5 | Payment or honoraria for lectures, presentations, speakers bureaus, manuscript writing or educational events | __x__ None |
| 6 | Payment for expert testimony | _x___ None |
| 7 | Support for attending meetings and/or travel | __x__ None |
| 8 | Patents planned, issued or pending | _x___ None |
| 9 | Participation on a Data Safety Monitoring Board or Advisory Board | __x__ None |
| 10 | Leadership or fiduciary role in other board, society, committee or advocacy group, paid or unpaid | __x__ None |
| 11 | Stock or stock options | _x___ None |
| 12 | Receipt of equipment, materials, drugs, medical writing, gifts or other services | __x__ None |
| 13 | Other financial or non-financial interests | __x__ None |

**Please place an "X" next to the following statement to indicate your agreement:**

**_x__ I certify that I have answered every question and have not altered the wording of any of the questions on this form.**

#### ICMJE DISCLOSURE FORM

**Date:17 June 2021__________________________________________________________**
**Your Name: Dr Chuin Kee____________________________________________________________**
**Manuscript Title: Clinical management and impact of scarlet fever in the modern era: findings from a cross-sectional study of cases in London, 2018-2019.**
**Manuscript number (if known):NA ________________________________________________**

**In the interest of transparency, we ask you to disclose all relationships/activities/interests listed below that are related to the content of your manuscript. "Related" means any relation with for-profit or not-for-profit third parties whose interests may be affected by the content of the manuscript. Disclosure represents a commitment to transparency and does not necessarily indicate a bias. If you are in doubt about whether to list a relationship/activity/interest, it is preferable that you do so.**

**The following questions apply to the author's relationships/activities/interests as they relate to the current manuscript only.**

**The author's relationships/activities/interests should be defined broadly. For example, if your manuscript pertains to the epidemiology of hypertension, you should declare all relationships with manufacturers of antihypertensive medication, even if that medication is not mentioned in the manuscript.**

**In item #1 below, report all support for the work reported in this manuscript without time limit. For all other items, the time frame for disclosure is the past 36 months.**

| | | **Name all entities with whom you have this relationship or indicate none (add rows as needed)** | **Specifications/Comments (e.g., if payments were made to you or to your institution)** |
|---|---|---|---|
| | | **Time frame: Since the initial planning of the work** | |
| 1 | All support for the present manuscript (e.g., funding, provision of study materials, medical writing, article processing charges, etc.) **No time limit for this item.** | ____ None | |
| | | **Time frame: past 36 months** | |
| 2 | Grants or contracts from any entity (if not indicated in item #1 above). | ____ None | |
| 3 | Royalties or licenses | ____ None | |

| | | |
|---|---|---|
| 4 | Consulting fees | ____ None |
| 5 | Payment or honoraria for lectures, presentations, speakers bureaus, manuscript writing or educational events | ____ None |
| 6 | Payment for expert testimony | ____ None |
| 7 | Support for attending meetings and/or travel | ____ None |
| 8 | Patents planned, issued or pending | ____ None |
| 9 | Participation on a Data Safety Monitoring Board or Advisory Board | ____ None |
| 10 | Leadership or fiduciary role in other board, society, committee or advocacy group, paid or unpaid | ____ None |
| 11 | Stock or stock options | ____ None |
| 12 | Receipt of equipment, materials, drugs, medical writing, gifts or other services | ____ None |
| 13 | Other financial or non-financial interests | ____ None |

**Please place an "X" next to the following statement to indicate your agreement:**

**_X__ I certify that I have answered every question and have not altered the wording of any of the questions on this form.**

#### ICMJE DISCLOSURE FORM

**Date:_24/6/2021________________________________________________________________________**
**Your Name: Sooria Balasegaram______________________________________________________________**
**Manuscript Title: <u>Clinical management and impact of scarlet fever in the modern era: findings from a cross-sectional study of cases in London, 2018-2019.</u>**
**Manuscript number (if known):_NA_____________________________________________________________**

**In the interest of transparency, we ask you to disclose all relationships/activities/interests listed below that are related to the content of your manuscript. "Related" means any relation with for-profit or not-for-profit third parties whose interests may be affected by the content of the manuscript. Disclosure represents a commitment to transparency and does not necessarily indicate a bias. If you are in doubt about whether to list a relationship/activity/interest, it is preferable that you do so.**

**The following questions apply to the author's relationships/activities/interests as they relate to the <u>current manuscript only</u>.**

**The author's relationships/activities/interests should be <u>defined broadly</u>. For example, if your manuscript pertains to the epidemiology of hypertension, you should declare all relationships with manufacturers of antihypertensive medication, even if that medication is not mentioned in the manuscript.**

**In item #1 below, report all support for the work reported in this manuscript without time limit. For all other items, the time frame for disclosure is the past 36 months.**

| | | **Name all entities with whom you have this relationship or indicate none (add rows as needed)** | **Specifications/Comments (e.g., if payments were made to you or to your institution)** |
|---|---|---|---|
| | | **Time frame: Since the initial planning of the work** | |
| 1 | All support for the present manuscript (e.g., funding, provision of study materials, medical writing, article processing charges, etc.) **No time limit for this item.** | ___X_None | |
| | | **Time frame: past 36 months** | |
| 2 | Grants or contracts from any entity (if not indicated in item #1 above). | __X__None | |
| 3 | Royalties or licenses | __X__None | |
| 4 | Consulting fees | __X__None | |

| | | |
|---|---|---|
| 5 | Payment or honoraria for lectures, presentations, speakers bureaus, manuscript writing or educational events | __X__None |
| 6 | Payment for expert testimony | __X__None |
| 7 | Support for attending meetings and/or travel | __X__None |
| 8 | Patents planned, issued or pending | __X__None |
| 9 | Participation on a Data Safety Monitoring Board or Advisory Board | __X__None |
| 10 | Leadership or fiduciary role in other board, society, committee or advocacy group, paid or unpaid | __X__None |
| 11 | Stock or stock options | _X___None |
| 12 | Receipt of equipment, materials, drugs, medical writing, gifts or other services | _X___None |
| 13 | Other financial or non-financial interests | __X__None |

**Please place an "X" next to the following statement to indicate your agreement:**

**__X_ I certify that I have answered every question and have not altered the wording of any of the questions on this form.**

#### ICMJE DISCLOSURE FORM

**Date:17 June 2021__________________________________________________________________**
**Your Name:___________SHIRANEE SRISKANDAN**
**Manuscript Title: Clinical management and impact of scarlet fever in the modern era: findings from a cross-sectional study of cases in London, 2018-2019.**
**Manuscript number (if known):NA __________________________________________________________**

**In the interest of transparency, we ask you to disclose all relationships/activities/interests listed below that are related to the content of your manuscript. "Related" means any relation with for-profit or not-for-profit third parties whose interests may be affected by the content of the manuscript. Disclosure represents a commitment to transparency and does not necessarily indicate a bias. If you are in doubt about whether to list a relationship/activity/interest, it is preferable that you do so.**

**The following questions apply to the author's relationships/activities/interests as they relate to the current manuscript only.**

**The author's relationships/activities/interests should be defined broadly. For example, if your manuscript pertains to the epidemiology of hypertension, you should declare all relationships with manufacturers of antihypertensive medication, even if that medication is not mentioned in the manuscript.**

**In item #1 below, report all support for the work reported in this manuscript without time limit. For all other items, the time frame for disclosure is the past 36 months.**

| | | **Name all entities with whom you have this relationship or indicate none (add rows as needed)** | **Specifications/Comments (e.g., if payments were made to you or to your institution)** |
|---|---|---|---|
| | | **Time frame: Since the initial planning of the work** | |
| 1 | All support for the present manuscript (e.g., funding, provision of study materials, medical writing, article processing charges, etc.) **No time limit for this item.** | Action Medical Research | Funding to Imperial College London to support research |
| | | UKRI | Funding to Imperial College London to support research and public involvement |
| | | **Time frame: past 36 months** | |
| 2 | Grants or contracts from any entity (if not indicated in item #1 above). | X None | |
| 3 | Royalties or licenses | X None | |

| | | |
|---|---|---|
| 4 | Consulting fees | X None |
| 5 | Payment or honoraria for lectures, presentations, speakers bureaus, manuscript writing or educational events | X None |
| 6 | Payment for expert testimony | X None |
| 7 | Support for attending meetings and/or travel | X None |
| 8 | Patents planned, issued or pending | X None |
| 9 | Participation on a Data Safety Monitoring Board or Advisory Board | X None |
| 10 | Leadership or fiduciary role in other board, society, committee or advocacy group, paid or unpaid | X None |
| 11 | Stock or stock options | X None |
| 12 | Receipt of equipment, materials, drugs, medical writing, gifts or other services | X None |
| 13 | Other financial or non-financial interests | X None |

**Please place an "X" next to the following statement to indicate your agreement:**

**__X_ I certify that I have answered every question and have not altered the wording of any of the questions on this form.**
